# Safety and Immunogenicity of a VLP Poliovirus Vaccine: A Phase 1 Trial

**DOI:** 10.64898/2026.08.14.26360426

**Authors:** Christina C. Chang, Ruijie Wang, Sumeyya Ahmed, Yujia Chen, Basit Jafri, Camilla Lucy Smith, Bernardo A. Mainou, Lina Wang, Xiaoyuan Zhao, Meixu Yan, Haitao Huang, Qiaoling Yan, Luis Barreto, Jinbo Gou, Tao Zhu

## Abstract

**BACKGROUND:** Current polio vaccines face challenges including vaccine-derived poliovirus and high-containment manufacturing. We evaluated a recombinant trivalent virus-like particle (VLP)-based poliovirus vaccine (VPV) for safety and immunogenicity in a first-in-human phase 1 trial.

**METHODS:** In this randomized, observer-blind, active-controlled trial, 72 healthy adults (18 to 54 years) were assigned (1:1:1:1) to receive a single dose of VPV at low (45:8:25 D-antigen units [DU] + 0.1 mg aluminum phosphate [AP]), medium (45:8:25 DU + 0.3 mg AP), or high (90:12:45 DU + 0.3 mg AP) doses, or conventional inactivated poliovirus vaccine (cIPV). Primary outcomes were safety and tolerability. Secondary outcomes included neutralizing antibody titers through day 180.

**RESULTS:** No serious adverse events or Grade 3 reactions were reported. Solicited adverse events were reported in 77.8%, 55.6%, and 72.2% of the low-, medium-, and high-dose VPV groups, respectively, and 66.7% in the cIPV group. By day 29, VPV induced dose-dependent neutralizing antibody responses. For serotypes 1 and 2, the high-dose VPV group achieved geometric mean titers (GMTs) of 73,582 (95% CI, 31,198-173,545) and 110,623 (95% CI, 59,276-206,451), respectively, comparable to cIPV at 45,161 (95% CI, 20,973-97,244) and 112,361 (95% CI, 58,280-216,625). Although serotype 3 GMTs were lower for the high-dose VPV at 18,905 (95% CI, 8737-40,906) than for cIPV at 61,431 (95% CI, 31,123-121,251), 100% of high-dose VPV recipients achieved neutralizing titers ≥1:1024.

**CONCLUSIONS:** A single dose of VPV was safe and highly immunogenic, supporting its potential as a next-generation vaccine to advance global polio eradication. (Funded by the Gates Foundation and Tianjin Leading Enterprises Innovative project 23YDLQSY00100; ClinicalTrials.gov number, NCT06101173).

---

Poliomyelitis was the leading cause of flaccid paralysis and permanent disability in children in the pre-vaccine era. The widespread use of trivalent conventional inactivated poliovirus vaccine (cIPV) and OPV propelled by the Global Polio Eradication Initiative (GPEI) launched in 1988 led to 99% reduction in the number of children paralyzed by polio. However, as of December 2025, wild poliovirus (WPV) transmission continues in Pakistan and Afghanistan, and outbreaks of variant poliovirus continue to be reported in 27 countries, underscoring the persistent and evolving challenges to global eradication efforts. Although the cheaper and easier to administer OPV and its variants (including mono-, bi-, tri-, and novel Sabin strains) have played a critical role in global eradication, vaccine virus excretion and transmission to secondary contacts may occur. In settings with suboptimal vaccination coverage, this can contribute to outbreaks of circulating vaccine-derived polioviruses (cVDPVs) and cases of vaccine-associated paralytic poliomyelitis (VAPP).^3–9^ Although genetically stabilized novel OPV2 (nOPV2) was developed to reduce the risk of reversion, its deployment has not fully eliminated the emergence of cVDPV2.^10,11^ IPV avoids the risk of genetic reversion and provides strong systemic immunity; however its production requires high-level biocontainment, contributing to higher costs and limited accessibility in low-resource settings.^12^ These challenges highlight the need for a next-generation vaccine platform that improves biosafety and manufacturing scalability while maintaining robust immunogenicity.

Virus-like particles (VLPs) technology offers a promising alternative to currently-used polio vaccines. These non-infectious, self-assembling structures mimic the native viral capsid but lack any viral genetic material, eliminating risks of VAPP and cVDPV. VLPs do not require high-containment production facilities, making them a cost-effective and scalable option.^6,7,13,14^ Native-like poliovirus VLPs have been variously produced in mammalian, insect, bacterial, and yeast systems.^12,13,15,16^ However, the prototype particles suffered from poor thermal stability impacting on vaccine potency. Advances in virology have introduced stabilizing mutations, creating genetically engineered VLPs that preserve immunogenic D-antigen conformation and improved thermostability, obviating the need for cold-chain.^17–18^

Building upon these advancements, CanSino Biologics Inc., developed the Recombinant Trivalent VLP-based Poliovirus Vaccine (VPV), incorporating stabilized capsid proteins from poliovirus serotypes 1, 2, and 3, expressed in Sf-RVN^®^ insect cells.^19^ To our knowledge, this is the first VLP-based polio vaccine to progress to clinical evaluation. Here, we report the safety, tolerability, and immunogenicity data from the first-in-human Phase 1 trial of this novel VLP-based vaccine, given as a single intramuscular dose, in three different formulations in healthy adults aged 18–54 years.

## METHODS

### TRIAL OVERSIGHT

This first-in-human, randomized, observer-blind, positive-controlled, phase 1 trial was conducted at a single center, Nucleus Network in Melbourne, Australia. The protocol and informed consent forms were approved by the Bellberry Human Research Ethics Committee on December 15, 2023 (NCT06101173). The trial adhered to the Declaration of Helsinki and the International Conference on Harmonization Good Clinical Practice (ICH-GCP) guidelines. A Safety Review Committee reviewed safety data throughout the study. The trial was sponsored by CanSino Biologics, which developed the protocol in collaboration with the investigators. The study was funded by Gates Foundation and Tianjin leading enterprises innovative project. Several authors are affiliated with the funder and contributed to the manuscript preparation. All authors had full access to the data and vouch for the accuracy and completeness of the data and for the fidelity of the trial to the protocol.

### PARTICIPANTS

Eligible healthy adults aged 18 to 54 years, in good general health or with stable, mild medical conditions not requiring treatment changes in the prior six months, provided written informed consent and were enrolled between January 15 and March 8, 2024. Key inclusion and exclusion criteria are outlined in the Supplementary Appendix.

### TRIAL PROCEDURES

Three sequential groups of 24 participants were enrolled and randomized in a 3:1 ratio within each group to receive either one of three formulations of VPV or an active positive control (cIPV, IPOL^®^, Sanofi Pasteur), all administered as a single 0.5 ml intramuscular injection. The VPV formulations varied in D-antigen units (DU) for Sabin serotype 1, Salk serotype 2, and Sabin serotype 3, and in their aluminum phosphate (AP) adjuvant content, namely low dose (45:8:25 DU + 0.1 mg AP), medium dose (45:8:25 DU + 0.3 mg AP), and high dose (90:12:45 DU + 0.3 mg AP) VPV formulations. The comparator cIPV contained 40:8:32 DU of antigen for serotype 1 (Mahoney), 2 (MEF-1), and 3 (Saukett), respectively.

Each group began with 4 sentinel participants followed by a 3-day safety observation before vaccinating the remaining 20 participants. The SRC reviewed the 7-day safety data before escalating to the next group. Six onsite visits occurred: screening visit, vaccination on Day 1, and follow-up visits on Day 3, 8, 29, and 180 post-vaccination. Participants were required to report solicited (local and systemic) and unsolicited adverse events (AEs) in a paper diary up to Day 29. Safety laboratory tests were performed on blood samples collected on Days 1, 3, and 8, while immunogenicity assessments for neutralizing antibodies were performed on blood samples collected on Day 29 and 180.

### OUTCOMES

The primary safety outcome was the incidence of solicited local and systemic AEs within 7 days post-vaccination. Secondary outcomes included AEs within 30 minutes post-vaccination, unsolicited AEs through Day 29 as recorded in diaries, laboratory abnormalities (hematology and biochemistry) assessed at screening and on Days 1, 3, and 8, and serious adverse events (SAEs) throughout the 6-month follow-up period.

Neutralizing antibody titers against Sabin poliovirus serotypes 1, 2, and 3 at baseline, Day 29, and Day 180 using microneutralization assays performed with HEp-2(C) cells according to WHO protocols performed by the United States Centers of disease control and prevention (CDC).^20^ Titers were reported as the reciprocal of the highest serum dilution that neutralized virus replication. Seroprotection for poliovirus was defined as neutralizing antibody titers ≥1:8, and seroconversion was defined as a fourfold rise in titers among seropositive participants or attainment of titers ≥1:8 in seronegative individuals.^21^ Geometric mean titers (GMTs) and geometric mean fold increases (GMIs) were calculated accordingly.

### STATISTICAL ANALYSIS

This was an exploratory phase 1 trial, and sample size was not powered for formal hypothesis testing. Safety data were analyzed descriptively. Incidence of AEs and laboratory abnormalities was compared between VPV and cIPV groups using Chi-square or Fisher’s exact tests, as appropriate. Immunogenicity was assessed in participants with both baseline and post-vaccination data. GMTs and 95% confidence intervals (CIs) were computed and compared using the Wilcoxon rank-sum test. Seroconversion rates were presented with 95% CIs using the Clopper-Pearson method. A two-sided P value ≤0.05 was considered statistically significant, with no adjustment for multiple testing. Analyses were performed using SAS version 9.4, and included the Intent-to-Treat (ITT), Safety (SS), Immunogenicity (IMM), and Per-Protocol (PPS) analysis sets.

## RESULTS

### CHARACTERISTICS OF THE PARTICIPANTS

Of the 158 individuals screened, 72 participants were enrolled and randomized into three VPV dose groups (low, medium, and high) and a cIPV control group. All participants received the assigned vaccine according to their group, and were included in the ITT, SS, and IMM analysis sets. Three participants (4.2%) discontinued early due to geographical relocation (Fig. 1). One participant in the high-dose VPV group was excluded from the PPS as their immunogenicity blood collection was out of protocol window. The cohort was majority female (59.7%), with a median age of 28.1 (IQR 19.8 to 54.3) years, and a median body mass index of 24.1 kg/m^2^. Most participants were White (47.2%) or Asian (37.5%), while 4 identified as Black or African American, 4 as American Indian or Alaska Native, and 3 as Other. Baseline demographics were similar across groups (Table S1 in Supplementary Appendix).

**Figure 1:**
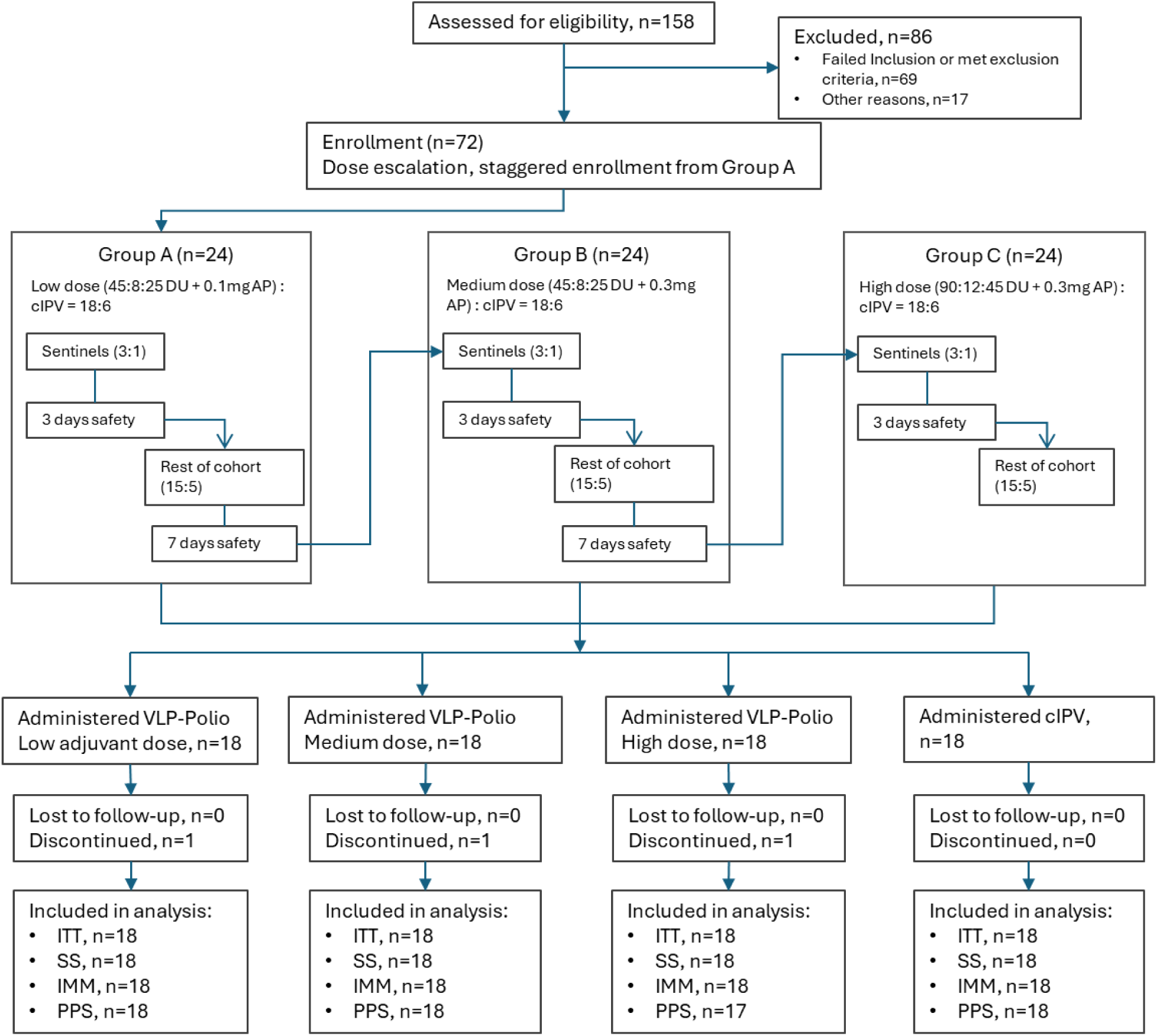
Study design of the Phase 1 trial. A total of 158 participants were assessed for eligibility, of whom 72 were enrolled and randomly assigned to receive low-, medium-, or high-dose VPV or cIPV in three sequential cohorts with dose escalation. In each cohort, sentinel participants (3 receiving VPV and 1 receiving cIPV) were observed for 3 days for safety before enrollment of the remaining participants, with additional safety evaluation through 7 days before dose escalation. All participants received a single intramuscular dose and were followed through Day 180. No participants were lost to follow-up; three discontinued. All participants completed assessments for primary safety endpoints and Day 29 immunogenicity analysis. ITT intention-to-treat, SS safety set, IMM immunogenicity set, and PPS per-protocol set.

### SAFETY

All vaccine formulations were generally well tolerated. Among the 18 participants in each group, solicited AEs within 7 days were reported in 14, 10, 13, and 12 participants in the low-, medium-, high-dose VPV, and cIPV groups, respectively (Table 1). The cIPV group had the highest ratio of Grade 2 to Grade 1 events. Grade 1 injection site pain was reported in 12, 8, 10, and 9 participants, and Grade 2 in 0, 1, 1, and 2 participants in the low-, medium-, high-dose VPV, and cIPV groups, respectively. Only one participant each in the medium- and high-dose VPV groups reported Grade 1 injection site swelling, with the latter also reporting with Grade 1 induration. No erythema was observed.

**Table 1.**
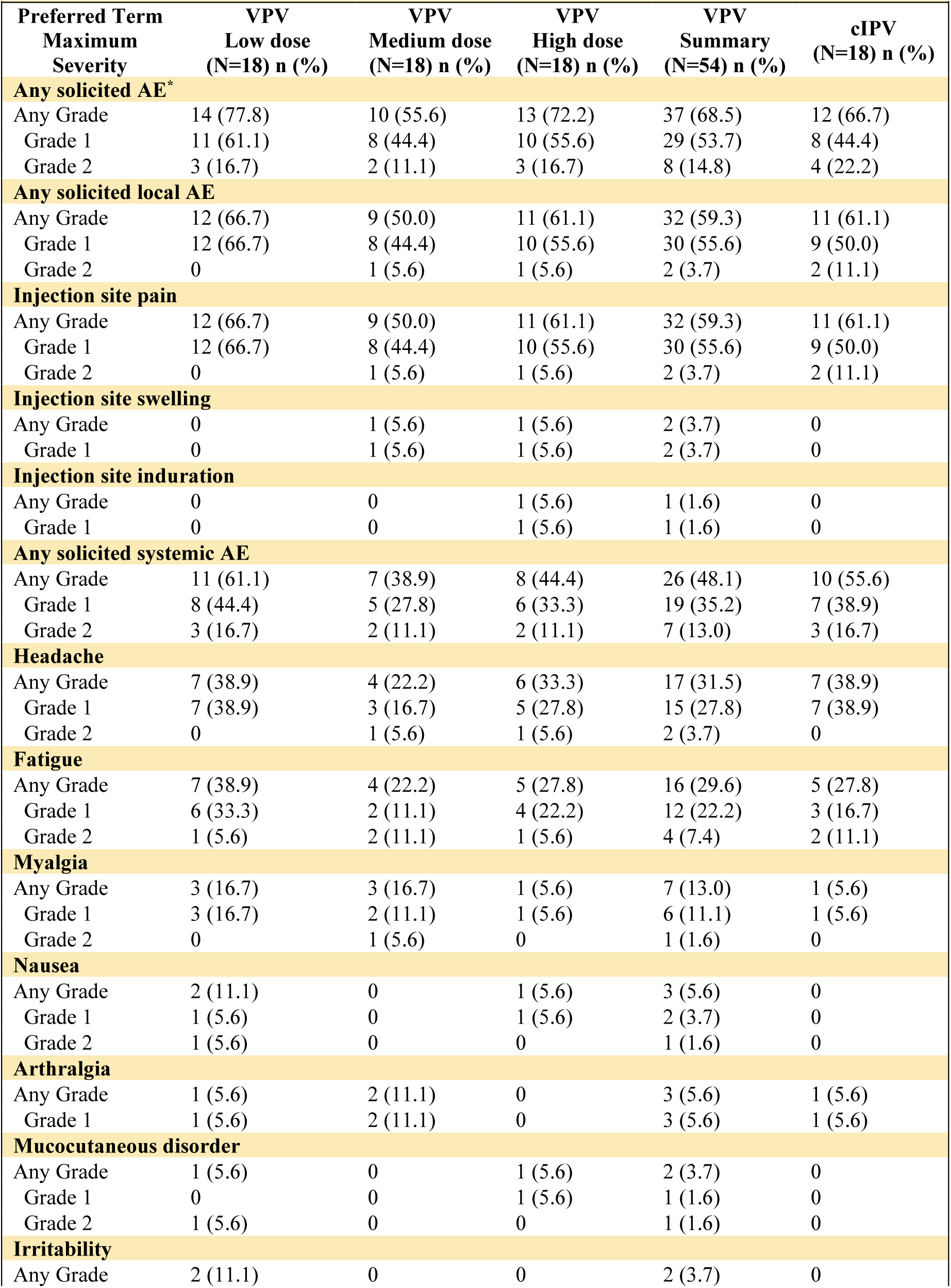
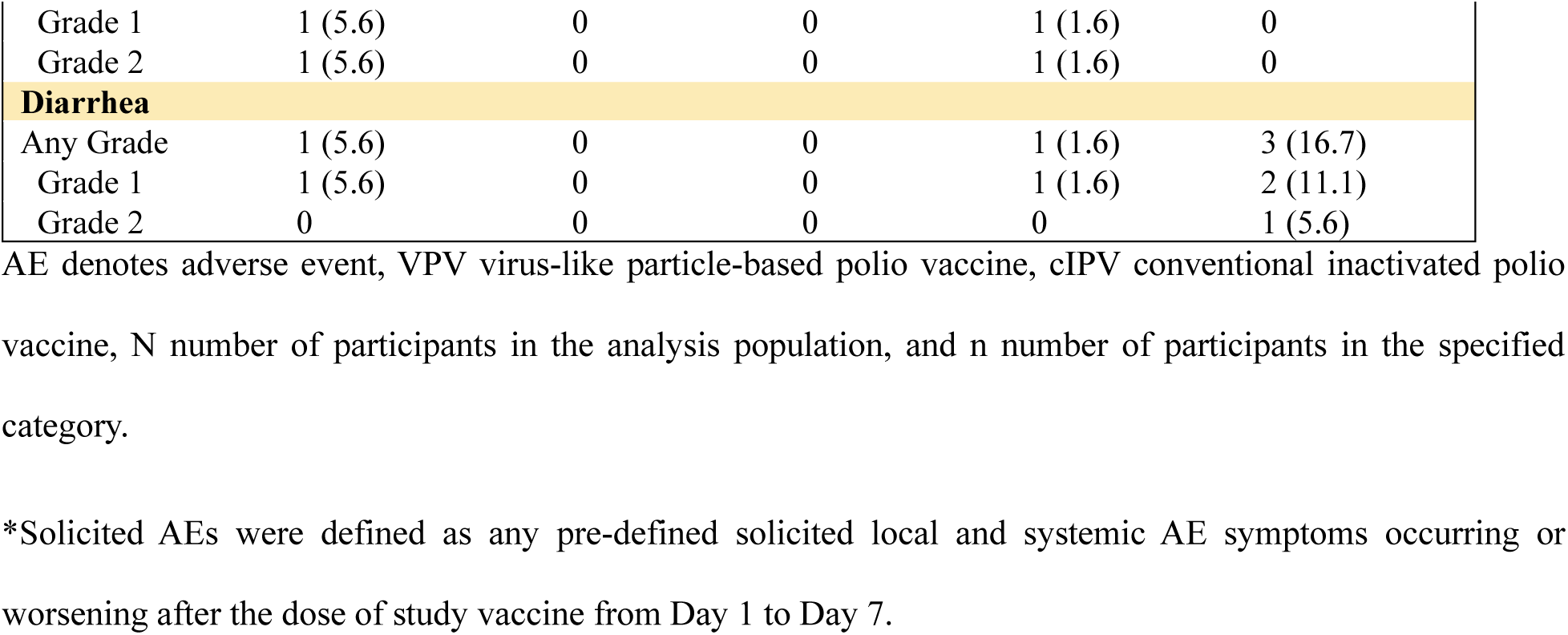
Summary of Solicited Adverse Events Within 7 Days Post-Vaccination.

Solicited systemic AEs were reported in 11, 7, 8, and 10 participants in the low-, medium-, high-dose VPV, and cIPV groups, respectively, with 72% of events classified as Grade 1. Headache and fatigue were the most common systemic AEs, reported in 7, 4, 6, and 7 participants, and 7, 4, 5, and 5 participants, in the low-, medium-, high-dose VPV, and cIPV groups, respectively, predominantly as Grade 1 events. No statistically significant differences in systemic AE rates were observed between groups. No fever, Grade 3 AEs, or SAEs were reported.

Unsolicited AEs through Day 28 were reported in 25% of participants, largely represented by viral respiratory tract infections, bowel habit changes and non-specific headaches. Most unsolicited AEs were considered unrelated to vaccination; four events (three in the medium-dose group and one in the high-dose group) were considered related (Table S2). Transient laboratory abnormalities were infrequent and mild. Transient decreases in neutrophil counts were observed in two (both Grade 1), one (Grade 2) and one (Grade 1) participant in the medium-, high-dose VPV, and cIPV groups, respectively. One participant in the cIPV group experienced a mild increase in creatinine (Table S3). All laboratory abnormalities were asymptomatic and did not require clinical intervention.

### IMMUNOGENICITY

Most participants exhibited baseline seroprotection (neutralizing antibody titer ≥1:8) prior to vaccination, with 93.1%, 92.7%, and 88.9% for serotype 1, 2, and 3, respectively. The overall mean GMT was 199 (95% CI, 123 to 322) and 241 (95% CI, 157 to 371) for serotype 1 and 2, and generally lower for serotype 3, with a GMT of 84 (95% CI, 49 to 143) (Table 2). No significant differences between vaccine groups at baseline. At Day 29, serotype 1 GMTs were highest in the high-dose group, reaching 73,582 (95% CI, 31,198 to 173,545), which was significantly greater than those in the medium-dose group (P*=*0.012), while remaining comparable to both the low-dose VPV and cIPV groups. For serotype 2, antibody responses were broadly similar across all VPV and cIPV groups, with GMTs of 71,743 (95% CI, 35,377 to 145,491), 54,058 (95% CI, 24,436 to 119,587), and 110,623 (95% CI, 59,276 to 206,451) for low-, medium-, and high-dose VPV groups, respectively, compared with 112,361 (95% CI, 58,280 to 216,625) for cIPV. In contrast, serotype 3 responses were substantially lower in all VPV groups compared with 61,431 (95% CI, 31,123 to 121,251) for cIPV. Specifically, GMTs were 18,165 (95% CI, 7,685 to 42,936; P*=*0.038), 15,369 (95% CI, 6,174 to 38,263; P*=*0.029), and 18,905 (95% CI, 8,737 to 40,906; P*=*0.023) in the low-, medium-, and high-dose VPV groups, respectively.

**Table 2.**
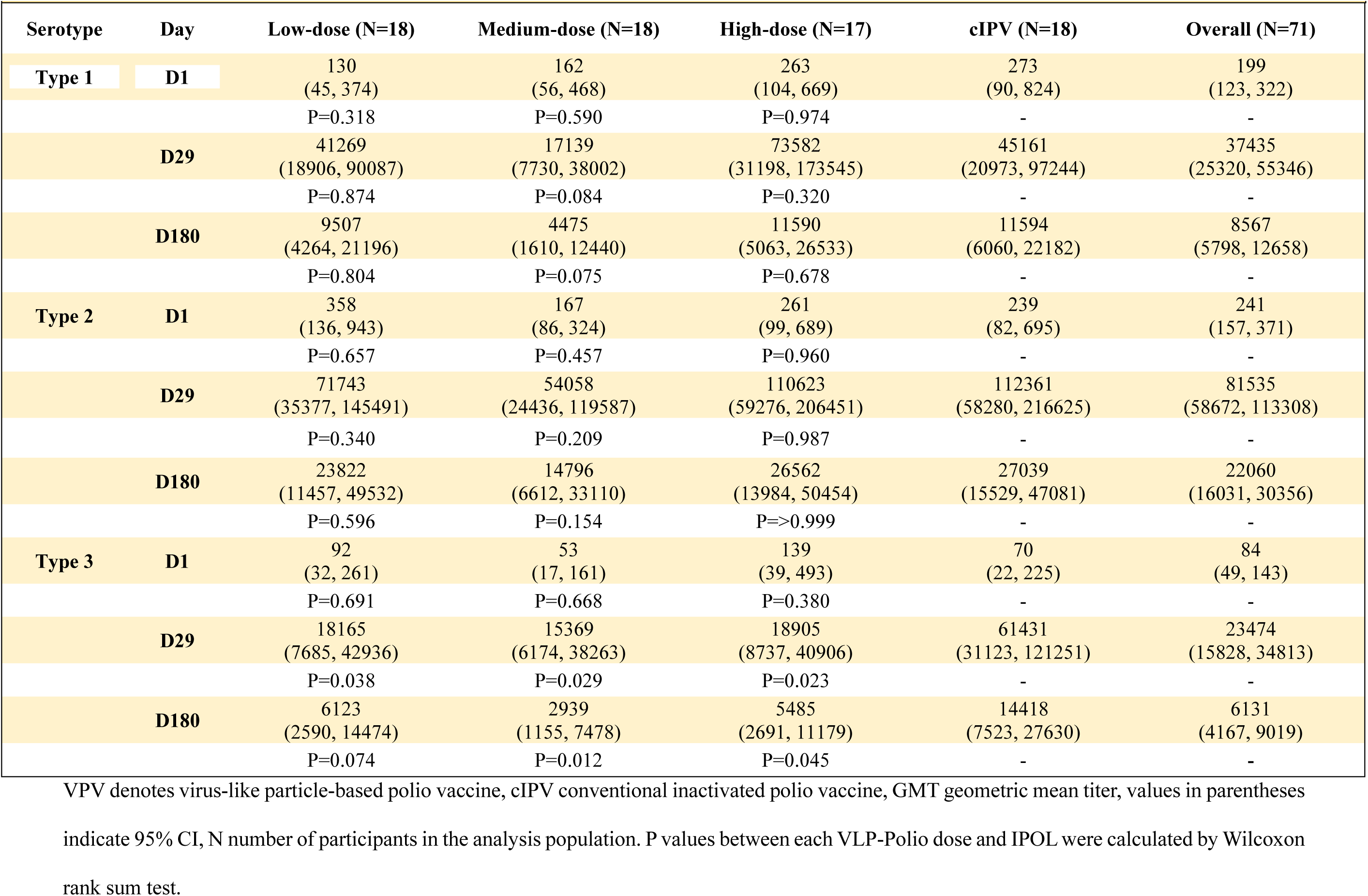
Neutralizing Antibody Responses (GMT) to Poliovirus Types 1, 2, and 3 at Baseline and Following Vaccination.

For serotype 1, GMIs at Day 29 were 318.2, 105.6, 279.5, and 165.5, and at Day 180 were 62.7, 25.9, 44.0, and 42.5 at Day 180, in the low-, medium-, high-dose VPV, and cIPV groups, respectively, with no significant difference between groups. For serotype 2, GMIs were 200.5, 324.7, 423.6, and 471.0 at Day 29 and 53.6, 85.7, 101.7, and 113.3 at Day 180 across the same groups. For serotype 3, GMIs were 198.1, 292.9, 136.2, and 877.1 at Day 29, and 56.7, 53.3, 39.5, and 205.9, at Day 180 in the low-, medium-, high-dose VPV, and cIPV groups, respectively. No significant differences in GMI were observed between VPV-groups and cIPV, except for a marginally lower Day 180 GMI for the high-dose VPV group (P*=*0.049) (Table S4).

In view of these significantly higher neutralizing antibody titers compared to the conventional seroprotection threshold of 1:8, reverse cumulative distribution plots (Fig. 2 and Table 3) were used to characterize responses across higher titer ranges. Baseline (Day 1) titers were uniformly low across all study groups. By Day 29, serotype 1 curves showed a marked rightward shift. All participants achieved titers ≥1:512 and over 94% exceeded 1:2048. The high-dose VPV and cIPV groups showed the most robust and durable responses, with more than 88% maintaining this level through Day 180. Responses to serotype 2 were universally strong with 100% participants reached 1:2048 by Day 29, and all participants in the high-dose VPV group and the cIPV group remain ≥1:2048 at Day 180, compared to 94.1% and 88.2% in the low- and medium-dose VPV groups, respectively. For serotype 3, nearly all participants reached Day 29 titers ≥1:512, with the medium-dose VPV group being the only exception (94.4%). Antibody titer ≥1:1024 were 100% in both high-dose VPV group and the cIPV group, and ≥1:2048 were observed in 100% of cIPV recipients and in 94%, 72%, and 94% of participants in the low-, medium-, and high-dose VPV groups, respectively. By Day 180, titers ≥1:2048 were retained in 100%, 76%, 64%, and 88% of participants, respectively.

**Figure 2:**
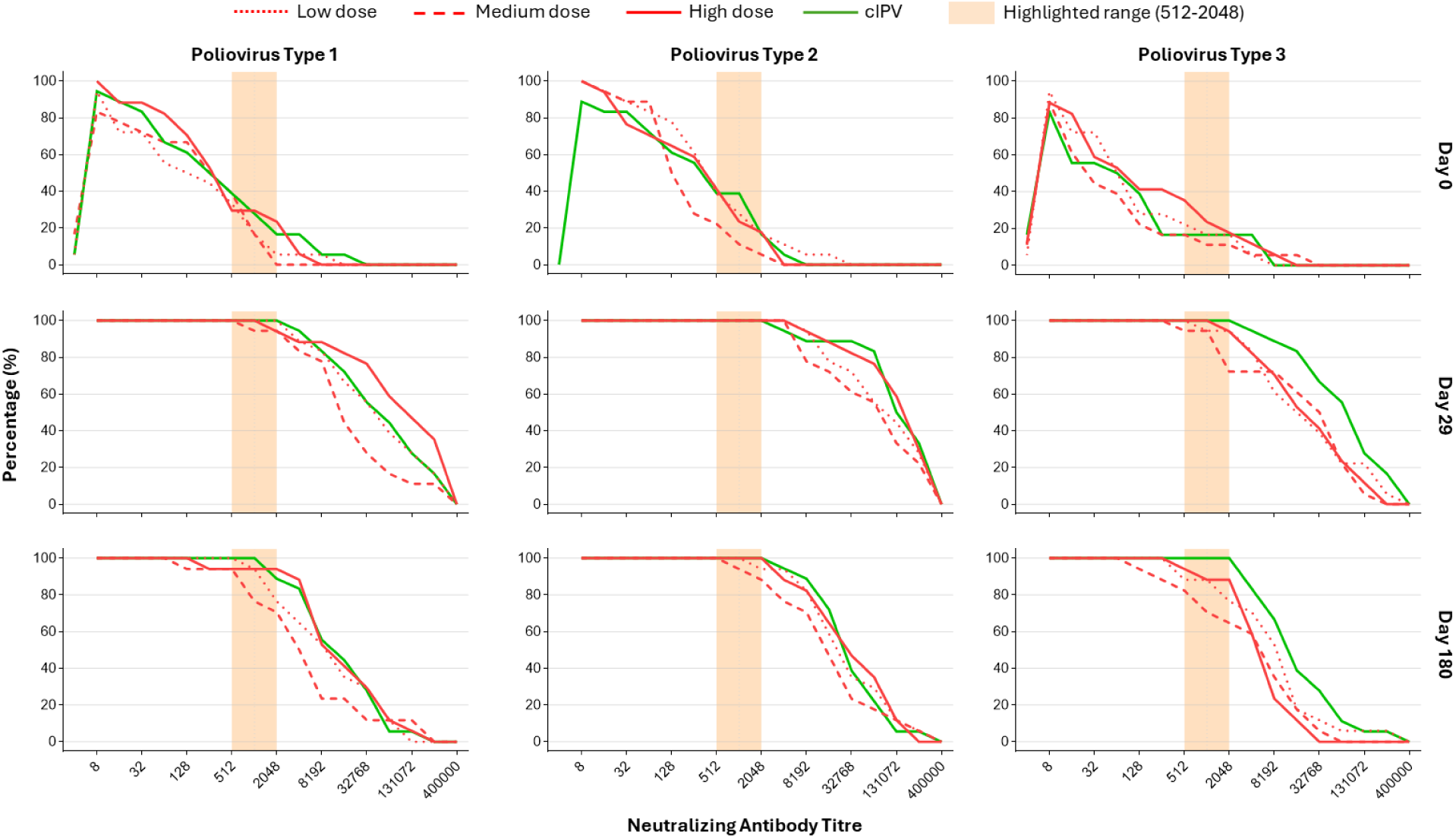
Cumulative antibody titer distribution curves at baseline, Day 29, and 180 for poliovirus type 1, type 2, and type 3. Cumulative distribution curves of neutralizing antibody titers against poliovirus types 1, 2, and 3 are shown at baseline and on days 29 and 180. Titers are expressed as the reciprocal of serum dilution. Low dose, medium dose, and high dose refer to VPV administered at three respective dosage levels, represented by the dotted red line, dashed red line, and solid red line. The solid green line denotes conventional inactivated poliovirus vaccine (cIPV). The shaded area indicates the prespecified protective range (512 to 2048).

**Table 3.** Seroconversion Rate and proportion of participants achieving various neutralizing antibody thresholds.

|  |  |  |  |  |  |  |  |  |  |  |  |  |  |
| --- | --- | --- | --- | --- | --- | --- | --- | --- | --- | --- | --- | --- | --- |
| Serotype | GMT threshold | Day 0 |  |  |  | Day 29 |  |  |  | Day 180 |  |  |  |
|  |  | Low dose (N=18) | Medium dose (N=18) | High dose (N=18) | cIPV (N=18) | Low dose (N=18) | Medium dose (N=18) | High dose (N=17) | cIPV (N=18) | Low dose (N=18) | Medium dose (N=18) | High dose (N=17) | cIPV (N=18) |
| Type 1 | ≥1:8 | 94.4% | 88.9% | 100.0% | 94.4% | 100.0% | 100.0% | 100.0% | 100.0% | 100.0% | 100.0% | 100.0% | 100.0% |
|  | ≥1:512 | 33.3% | 38.9% | 27.8% | 38.9% | 100.0% | 100.0% | 100.0% | 100.0% | 100.0% | 94.1% | 94.4% | 100.0% |
|  | ≥1:1024 | 16.7% | 16.7% | 1.5% | 27.8% | 100.0% | 94.4% | 100.0% | 100.0% | 94.1% | 76.5% | 94.4% | 100.0% |
|  | ≥1:2048 | 5.6% | 11.1% | 22.2% | 16.7% | 100.0% | 94.4% | 94.4% | 100.0% | 76.5% | 70.6% | 94.1% | 88.9% |
| Type 2 | ≥1:8 | 100.0% | 100.0% | 100.0% | 88.9% | 100.0% | 100.0% | 100.0% | 100.0% | 100.0% | 100.0% | 100.0% | 100.0% |
|  | ≥1:512 | 38.9% | 22.2% | 38.9% | 38.9% | 100.0% | 100.0% | 100.0% | 100.0% | 100.0% | 100.0% | 100.0% | 100.0% |
|  | ≥1:1024 | 27.8% | 11.1% | 22.2% | 38.9% | 100.0% | 100.0% | 100.0% | 100.0% | 100.0% | 94.1% | 100.0% | 100.0% |
|  | ≥1:2048 | 16.7% | 5.6% | 16.7% | 16.7% | 100.0% | 100.0% | 100.0% | 100.0% | 94.1% | 88.2% | 100.0% | 100.0% |
| Type 3 | ≥1:8 | 83.3% | 94.4% | 88.9% | 88.9% | 100.0% | 100.0% | 100.0% | 100.0% | 100.0% | 100.0% | 100.0% | 100.0% |
|  | ≥1:512 | 22.2% | 16.7% | 33.3% | 16.7% | 100.0% | 94.4% | 100.0% | 100.0% | 88.2% | 82.4% | 94.4% | 100.0% |
|  | ≥1:1024 | 16.7% | 11.1% | 22.2% | 16.7% | 94.4% | 94.4% | 100.0% | 100.0% | 88.2% | 70.6% | 88.9% | 100.0% |
|  | ≥1:2048 | 16.7% | 11.1% | 16.7% | 16.7% | 94.4% | 72.2% | 94.4% | 100.0% | 76.5% | 64.7% | 88.2% | 100.0% |

Seroprotection was defined as antibody level $\geq 1:8$ . Different GMT thresholds ( $\geq 1:512$ , $\geq 1:1024$ , and $\geq 1:2048$ ) were used to evaluate the proportion of participants after vaccination.

## DISCUSSION

In this first-in-human phase 1 clinical trial, a novel trivalent VLP-based poliovirus vaccine adjuvanted with AP and administered as a single intramuscular dose was as well tolerated as cIPV in adults aged 18-54 years. Across all dose levels, no increase in frequency or severity of solicited or unsolicited AEs was observed compared with cIPV.

Using the conventional definition of ≥1:8, baseline seroprotection rates were high (>88.9%) despite the relatively low corresponding antibody titers. All three VPV formulations, 45:8:25 DU with 0.1 mg or 0.3 mg AP adjuvant and 90:12:45 DU with 0.3 mg AP adjuvant, induced notable immune responses, characterized by >100 GMI at Day 29 (Table S4). The resulting GMT reached at least 17,139, 54,058, and 15,369 for serotypes 1, 2, and 3, respectively, which were all remarkably higher than 8. We found relatively low serotype 3 responses in the VPV groups compared to cIPV, potentially reflecting subtle antigenic divergence from the cIPV used for primary immunization, resulting in a less potent heterologous boost. Since serum neutralizing antibodies are the established correlate of protection against poliomyelitis, these robust immunogenicity profiles, characterized by seroprotection rates and antibody magnitudes comparable to the licensed cIPV, suggest potential clinical efficacy of the investigational product.^21^

The low- and medium-dose VPV formulations shared an identical antigen ratio of 45:8:25 DU but differed in AP adjuvant. This data suggested that that increasing the adjuvant dose from 0.1 to 0.3 mg AP provided no advantage in GMTs post-vaccination. However, higher AP adjuvant was associated with increased GMI for serotype 2 responses at Day 29 and 180, and for serotype 3 at Day 29. For serotype 3, 100% and 94% of participants in the low- and medium-dose VPV groups achieved titers of ≥1:512 at Day 29, declining to 88% and 82%, respectively, at 6 months.

Comparing the medium- to the high-dose VPV arms, where the former contained 45:8:25 DU and the latter nearly doubling this at 90:12:45 DU, both with the same 0.3 mg AP adjuvant, GMTs were markedly higher across all serotypes in the high-dose VPV group at Day 29 and 180. The GMI in the high-dose VPV group was higher than the medium group for serotypes 1 and 2, but not serotype 3. Importantly, at Day 29, 100% of participants in the high-dose VPV group achieved neutralizing antibodies of ≥1:512 compared to 94% in the medium-dose VPV group. All participants in the high group achieved titers of at least 1:1024 at Day 29 and 1:256 at Day 180, compared with 1:256 and 1:64, respectively, in the medium group. Solicited and unsolicited AEs were not more frequent nor more severe in the high-dose VPV group. Together, this suggests formulations containing higher-D-antigen units leads to more immunogenic response without compromising on safety. The increased AP adjuvant content seems to not improve immunogenicity by GMT nor its durability to Day 180.

This VLP based poliovirus vaccine tested in humans marks a major advancement in polio vaccine innovation, particularly relevant in the post-polio eradication era. This vaccine is produced using recombinant baculoviruses that express structural proteins from each poliovirus serotype 1, 2, and 3, introduced into insect Sf-RVN^®^ cell lines where they are cultured, expressed, harvested, purified and blended into specific DU proportions. Production is rapid and relatively low cost. This VPV vaccine has no preservatives or antibiotics and can be stored at 2-8°C and remains stable for 24 months. Importantly, because this vaccine contains no live virus or genetic material, there is no risk for vaccine-derived polio variants. This VPV may be developed as a stand-alone vaccine as a primary series or as a booster, and may be formulated with other childhood vaccines as a combination vaccine.

Ongoing sporadic VAPP cases, outbreaks of cVDPV with OPV-like isolates and VDPV isolates with higher VP1 sequence divergence from their parental OPV strains is compelling an earlier global cessation of the OPV vaccine. The switch from trivalent OPV to bivalent OPV removing serotype 2 in response to VDPV2 outbreaks recommended by the WHO, together with vaccine supply constraints and lower vaccine coverage, has led to large populations of children without seroprotection to serotype 2.^22^ Considerable efforts are underway to optimize the timing and frequency of primary immunization series in polio prevention, using a mix of OPV and IPV, adjusting dosing regimen, and/or optimizing IPV immunogenicity and improving its production costs with adjuvants, novel strains, and varying antigen doses. Development of next-generation polio vaccines are therefore urgently needed to break the final frontier of global polio eradication and to ensure long-term protection in the post-eradication era.

The favorable safety and immunogenicity profile observed in this study supports the continued clinical development of this VLP-based poliovirus vaccine in larger trials in diverse populations, particularly infants and toddlers, to confirm these findings and to evaluate different vaccination schedules. In conclusion, this first-in-human trial of a trivalent VLP-based poliovirus vaccine demonstrated a favorable safety profile and strong immunogenicity in adults. These findings represent a meaningful step forward in the development of next-generation polio vaccines, with the potential to support the long-term goal of a polio-free world.

This work was supported, in whole or in part, by the Gates Foundation [INV-064409]. The conclusions and opinions expressed in this work are those of the author(s) alone and shall not be attributed to the Foundation. Under the grant conditions of the Foundation, a Creative Commons Attribution 4.0 License has already been assigned to the Author Accepted Manuscript version that might arise from this submission.

Disclosure forms provided by the authors are available with the full text of this article at NEJM.org.

Deidentified individual participant data that underlie the results reported in this article will be made available upon reasonable request to CanSino Biologics, subject to review and approval. A data access agreement will be required.

We acknowledge support from the Tianjin Leading Enterprises Innovative project 23YDLQSY00100 (Tianjin, China). We thank David Vaughn, Anna Du, Rachel Burke, Ruvim Izikson, and Peter Dull of Gates Foundation for providing insights throughout the study design and trial implementation. We are grateful to all participants and their families for their participation in the trial; members of the safety review committee for their review of the data; and Edwin Wong from Nucleus Network for coordinating the trial; and Rebecca Playne and Roopa Konanur from Novotech CRO (Melbourne, Australia) for medical-writing and study management assistance.

## Data Availability

All data produced in the present study are available upon reasonable request to the authors.

## REFERENCES

1 Tang X, Xiao Y, Deng X, et al. Immuno-persistence of the different primary polio vaccine schedules and immunogenicity of the booster dose by sabin inactivated or bivalent oral poliovirus vaccine in children aged 4 years: an open-label, randomised, controlled phase 4 trial in China. Lancet Reg Health West Pac 2023 17; 34:100725.

2 Global Polio Eradication Initiative. Wild poliovirus list. Polio Eradication; 2022. Accessed April 7, 2026. https://polioeradication.org/polio-today/polio-now/wild-poliovirus-list/

3 Bashorun AO, Kotei L, Jawla O, et al. Tolerability, safety, and immunogenicity of the novel oral polio vaccine type 2 in children aged 6 weeks to 59 months in an outbreak response campaign in The Gambia: an observational cohort study. Lancet Infect Dis 2024; 24:417–26.

4 World Health Organization. Global wild poliovirus 2016–2022. World Health Organization; August 2022. Accessed April 7, 2026. https://polioeradication.org/wp-content/uploads/2022/09/weekly-polio-analysesWPV-2022-20220830.pdf

5 Walter K, Malani PN. What is polio? JAMA. 2022; 328:1652.

6 Modlin JF, Bandyopadhyay AS, Sutter R. Immunization Against Poliomyelitis and the Challenges to Worldwide Poliomyelitis Eradication. J Infect Dis 2021; 224: S398–S404

7 Quarleri J. Poliomyelitis is a current challenge: long-term sequelae and circulating vaccine-derived poliovirus. Geroscience 2023; 45:707–17.

8 Voorman A, Lyons H, Bennette C, Kovacs S, Makam JK, F Vertefeuille J, Tallis G. Analysis of population immunity to poliovirus following cessation of trivalent oral polio vaccine. Vaccine 2023; 41:A85–92.

9 Macklin GR, O’Reilly KM, Grassly NC, et al. Evolving epidemiology of poliovirus serotype 2 following withdrawal of the serotype 2 oral poliovirus vaccine. Science 2020; 368:401–05.

10 Yeh MT, Bujaki E, Dolan PT, et al. Engineering the live-attenuated polio vaccine to prevent reversion to virulence. Cell Host Microbe 2020; 27:736–51.

11 Chumakov K, Ehrenfeld E, Agol VI, Wimmer E. Polio eradication at the crossroads. Lancet Glob Health 2021; 9:e1172–75.

12 Mbani CJ, Nekoua MP, Moukassa D, Hober D. The Fight against Poliovirus Is Not Over. Microorganisms 2023; 11:1323.

13 Sherry L, Grehan K, Swanson JJ, et al. Production and Characterisation of Stabilised PV-3 Virus-like Particles Using Pichia pastoris. Viruses 2022; 14: 2159.

14 World Health Organization. Polio Eradication Strategy 2022-2-26: Delivering on a Promise. Geneva, Switzerland: World Health Organization; 2021.

15 Bahar MW, Porta C, Fox H, Macadam AJ, Fry EE, Stuart DI. Mammalian expression of virus-like particles as a proof of principle for next generation polio vaccines. NPJ Vaccines 2021; 6:5.

16 Marsian J, Fox H, Bahar MW, Kotecha A, Fry EE, Stuart DI, Macadam AJ, Rowlands DJ, Lomonossoff GP. Plant-made polio type 3 stabilized VLPs-a candidate synthetic polio vaccine. Nat Commun 2017; 8:245.

17 Xu Y, Ma S, Huang Y, et al. Virus-like particle vaccines for poliovirus types 1, 2, and 3 with enhanced thermostability expressed in insect cells. Vaccine 2019; 37:2340–47.

18 Sherry L, Bahar MW, Porta C, et al. Recombinant expression systems for production of stabilized virus-like particles as next generation polio vaccines. Nat Commun 2025; 16:831.

19 Maghodia AB, Geisler C, Jarvis DL. Characterization of an Sf-rhabdovirus-negative Spodoptera frugiperda cell line as an alternative host for recombinant protein production in the baculovirus-insect cell system. Protein Expr Purif. 2016; 122:45–55.

20 Weldon WC, Oberste MS, Pallansch MA. Standardized methods for detection of poliovirus antibodies. Methods Mol Biol. 2016; 1387:145–176.

21 Expert Committee on Biological Standardization. Recommendations to assure the quality, safety and efficacy of poliomyelitis vaccines (oral, live, attenuated). In: WHO Expert Committee on Biological Standardization: sixty-third report. Geneva: World Health Organization; 2014: 83-172. (WHO technical report series, no. 980)

22 Macklin GR, O’Reilly KM, Grassly NC, et al. Evolving epidemiology of poliovirus serotype 2 following withdrawal of the serotype 2 oral poliovirus vaccine. Science 2020; 368(6489): 401–405.

